# Survival analysis methods for analysis of hospitalization data: Application to COVID-19 patient hospitalization experience

**DOI:** 10.1101/2021.04.14.21255511

**Authors:** Paul J. Rathouz, Victoria Valencia, Patrick Chang, David Morton, Haoxiang Yang, Ozge Surer, Spencer Fox, Lauren Ancel Meyers, Elizabeth C. Matsui, Alex B. Haynes

## Abstract

During most of 2020, the COVID-19 pandemic gave rise to considerable and growing numbers of hospitalizations across most of the U.S. Typical COVID-19 hospitalization data, including length of stay, intensive care unit (ICU) use, mechanical ventilation (Vent), and in-hospital mortality provide clearly interpretable health care endpoints that can be compared across population strata. They capture the resources consumed for the care of COVID-19 patients, and analysis of these endpoints can be used for resource planning at the local level. Yet, hospitalization data embody novel features that require careful statistical treatment to be useful in this context. Specifically, statistical models must meet three goals: (i) They should mesh with and inform mathematical epidemiologic or agent-based models of the COVID-19 experience in the population. (ii) They need to handle administrative censoring of hospitalization experience when data are extracted and downloaded for a given patient before that patient’s hospitalization experience has terminated. And, (iii) models need to handle risks for competing events, the occurrence of one blocking the possibility of the other(s). For example, live discharge from the hospital “competes with” (i.e., blocks) in-hospital mortality. We have adapted approaches from the survival analysis literature to address these challenges in order to better understand and quantify the population experience in hospital with respect to length of stay, ICU, Vent use and so on. Using hospitalization data from a large U.S. metropolitan region, in this report, we show how standard techniques from survival analysis can be brought to bear to address these challenges and yield interpretable results. In the breakout/discussion, we will discuss formulation, estimation and inference, and interpretation of competing risks models.

## 1 Introduction

With a few exceptions, across most of the U.S., the COVID-19 pandemic began giving rise to considerable and growing numbers of hospitalizations during March, 2020 (Dong, 2021). Briefly, typical COVID-19 hospitalization, including length of stay, intensive care unit (ICU) use, mechanical ventilation (Vent), and in-hospital mortality are clear and comparable health care endpoints that can be compared across population strata. They capture the resources consumed for the care of COVID-19 patients, and analysis of these endpoints can be used for resource planning at the local level (Yang et al., 2020). In addition, parameters estimated from analysis of these endpoints can and have been used as inputs to mathematical epidemiological models (Wang et al., 2020).

Hospitalization data embody novel features that have required careful statistical treatment, approaches that we have adapted from the survival analysis methodological literature, developed, and applied since the early days of the pandemic in order to better understand and quantify the population experience in hospital with respect to length of stay, ICU, Vent use and so on. The models presented here have helped to bridge the statistical gap between agent-based epidemic models and time-to-event analysis in the presence of both competing risks and censoring. The purpose of this report is twofold. Primarily, we document and describe these statistical approaches, and illustrate illustrate them on an analysis from Summer, 2020, that informed modeling efforts by other members of our local team. Secondarily, we provide details on data exploited from one of three large hospital systems in a large metropolitan region of the United States.

To accomplish this first purpose, statistical models must first be parameterized such that they inform mathematical epidemiologic or agent-based models. Owing to the specification of such models both in terms of alternative pathways through disease “states” (e.g., susceptible, infectious, recovered) and in terms of length of time spent in various states, they are often framed “conditional on the future”. For example, models may be parameterized in terms of length of hospitalization stay “for those who die in hospital”, that is, conditional on the *future event* of dying in hospital. Statistical analysis of data informing such models will be more useful if such analyses are framed in these same terms. Additional challenges here also arise in several classic areas of biostatistical application; these include *censoring* and *competing risks*. In analysis of data on hospitalization and ICU use, *censoring* will arise when the data are administratively frozen and extracted for analysis when, for a subset of patients, the censoring time occurs before the event of interest. For example, when interest lies in the time from hospital admission to ICU admission, ICU admission for a given patient will be censored if the data are extracted for analysis while that patient is still in the hospital and has not yet experienced ICU admission. In fact, we do not know based on the data whether s/he will ever experience ICU admission or, in the alternative, if s/he will exit the hospital without accessing the ICU at all. Lastly, a related but trickier phenomenon is that of *competing risks* (Prentice et al., 1978; Gray, 1988; Fine & Gray, 1999). The problem is easiest to understand in terms of live discharge and in-hospital death: As a patient advances through time, s/he eventually will be discharged alive or will die in hospital. (Exactly) one of the two events must occur, and they cannot both occur. The two events are “competing” as to which one will occur first and will thereby block the other from occurring.

The approach and analyses presented here were motivated by nearly complete COVID-19 hospitalization data extracted from the electronic health record (EHR) system of one of three multi-hospital systems in a large metropolitan area in the Southwestern region of the United States, Austin, Texas. In the context of expanding data and ongoing analysis needs for purposes of modeling and local epidemic planning, we produced multiple instances of the analyses developed and illustrated here. At this point, the data are somewhat outdated. Our main goal here, however, is to use them as a platform to describe, document, and illustrate the statistical approach; more up-to-date and in-depth analyses are reported elsewhere (Rathouz, 2021). A second goal is to tabulate the estimates used in a modeling report from our group which describes a staged alert system designed to ensure healthcare capacity in the Central Texas region (Yang, 2020). A third goal is to provide some technical details on the procedures and variables involved in data extraction.

In Section (2), we describe in mathematical terms the data configuration, notation, and statistical models. The example data are described and analyzed using these approaches for illustrative purposes in Section (3), and a brief Discussion appears in Section (4).

## 2 Statistical Framework

### 2.1 Notation and Data Configuration

For a given hospitalization, let *t*_0_ be the admission date, *t*_ICU_ be the date of ICU admission or the date of a competing event—but not a censoring event—for ICU admission, *t*_Vent_ be a similar time for date of first use of mechanical ventilation, and *t*_Disch_ be the date of discharge or in-hospital death, whichever comes first. Denote a separate censoring date as *t*_Cens_. Note importantly that censoring does not block any of the events from actually occurring; it just masks our ability to observe that event in our data.

We want to define durations and also acknowledge that we can only observe the first of a set of competing events. As such define time durations in hospital for these events as:

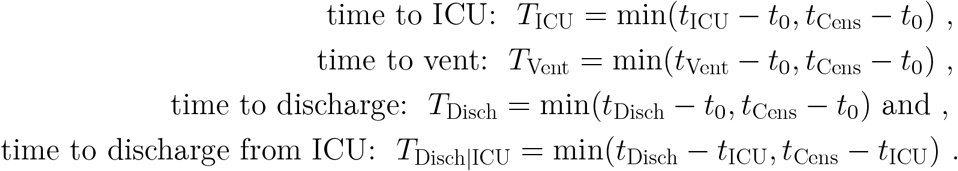

The last value *T*_Disch|ICU_ is only defined for subjects who are admitted to ICU, i.e., for whom *D*_ICU_ = 1, defined next.

Together with the various event times *t* and durations *T*, define corresponding event types *D*. For example, for ICU admission, *D*_ICU_ ∈ {0, 1, 2} indicates censoring while the patient is still in hospital and not-yet admitted to ICU (0), discharge from or deceased in hospital before any ICU admission (2), or admitted to ICU (1); and similarly for *D*_Vent_. Define *D*_Disch_ ∈ {0, 1, 2} to indicate censoring while the person is still in hospital, death in hospital (2), or live discharge from hospital (1). Note that under this definition, *T*_ICU_ = *t*_ICU_ − *t*_0_ when *D*_ICU_ ≥ 1 and *T*_ICU_ = *t*_Cens_ − *t*_0_ when *D*_ICU_ = 0, and similarly for *D*_Vent_ and *D*_Disch_. Two of these three event times, *T*_ICU_ and *T*_Disch|ICU_, and their corresponding event types, *D*_ICU_ and *D*_Disch_, are of primary interest in the analyses that follow, so we will focus on them. Finally, denote by *X* a vector of predictor or covariate values, which may include admission date, *t*_0_, may just be a stratifying variable, may be a regressor, or may not be included at all.

We frame these analyses in term of competing risks models (Prentice, 1978; Gray, 1988; Fine, 1999). Identifying assumptions in these models involve censoring time being conditionally independent of time to event of interest, given *X*. In our case, this is codified by

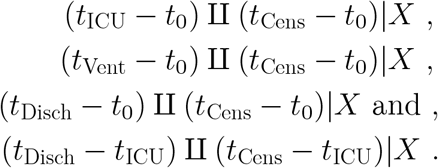

These assumptions would appear to be well justified by the nature of the problem, namely that administrative censoring for data extraction and download, is due only to processes determining when the facility pulls data from of their electronic health record and reports it to our team and/or to the Austin Public Health office.

### 2.2 Competing Risk Model Specification and Estimation

For any one of the foregoing time-to-event definitions (*T*_ICU_, *T*_Vent_, etc.) above, we consider a competing risks modeling framework. The data for each person and each event type *E* (e.g., *E* = ICU as defined above) consist of (*X, T*_*E*_, *D*_*E*_), where *X* and *T*_*E*_ are as defined in the foregoing section, and *D*_*E*_ = 0, 1,…, *K*_*E*_, where *D*_*E*_ = 0 denotes independent censoring, *D*_*E*_ = *k, k* = 1,…, *K*_*E*_ indicate *K*_*E*_ different competing endpoints for the event *E* of interest (e.g., live discharge or in-hospital mortality for *D*_Disch_), and, when appropriate, *D*_*E*_ = 1 indicates the event of primary interest.

The competing risks framework is built up through a collection of sub-distribution, or cumulative incidence, functions (CIF). We have already defined independent censoring date *t*_Cens_; consider the counterfactual situation wherein *t*_Cens_ = ∞, so that every subject is observed until one of the competing events occurs. In that case, *D*_*E*_ *>* 0 for all subjects and all events; denote this counterfactual *D*_*E*_ as 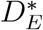. Then, the sub-distribution functions are defined as follows, where we omit the *E* subscript from *F, S, µ*, and *h*.

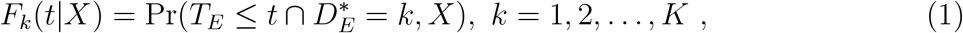

so that the overall distribution and survival functions for time-to-event endpoints are given by

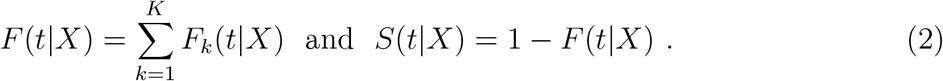

Note that these functions govern the distributions that would have been observed in the absence of independent administrative censoring. Estimation methods (Gray, 1988; Fine 1999) employed here account for censoring, yielding model interpretations in terms of (1) and (2), even while working with data involving such censoring, i.e., when *D*_*E*_—and not 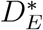—is observed.

An important feature of this formulation, and one that connects to the mathematical epidemic modeling framework, is that, as *t* → ∞, (1) yields the probability of failure being of type *k*, i.e.,

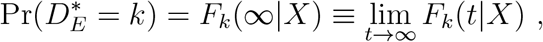

and in addition

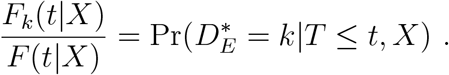

This means that, when analyzing times to events in hospitalized patients, we can also estimate the overall probability 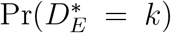 of event *k* occurring. Indeed, mathematical epidemic models such as Wang et al. (2020) are parameterized in terms of the hazard of event *k* occurring at some time *t, given* that it will eventually occur, i.e., that eventual failure will be of type *k*.

This hazard is sometimes called the instantaneous risk, or the failure *rate* in exponential waiting time models. When a given event type is of interest, the conditional (on event type *k*) distribution and hazard function are then given by

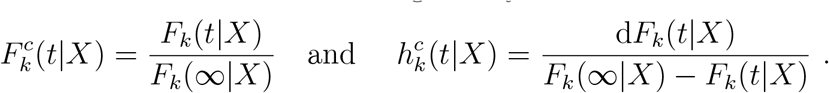

In the analyses presented here, key quantities to be estimated are *F*_*k*_(∞|*X*) as are features such as the mean and percentiles of the conditional distribution 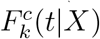.

In much of our work, covariate *X* is primarily used as a stratifying variable, and completely separate sets of CIF’s *F*_*k*_(*t*|*X* = *x*) are estimated for each value of *x*. For any given stratum *x*, we follow Gray (1988) to obtain estimates 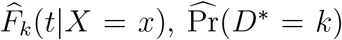, and 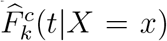, from which we can further obtain estimated conditional (on event type *k* occurring) quantiles 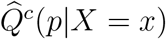 (where *Q*(·) is the inverse of *F* (·)), and the conditional mean failure time. This last quantity is conditional on event type *k* occurring and on stratum *x*; it is given by

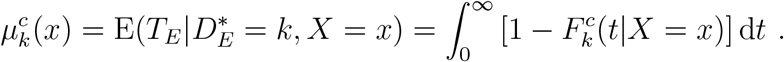

An alternative formulation treats *X* as a, possibly continuous and possibly vector-valued, covariate. In this case, analysis turns on the sub-distribution hazard function

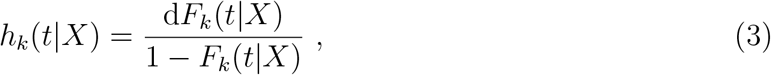

which is the instantaneous risk of event *k* at time *t* given event *k has not yet occurred* (even if another event *k*^*′*^ occurrence has blocked event *k*. Note that *h*_*k*_(*t*|*X*) is subtly different than the cause-specific hazard, which, loosely, is the instantaneous risk of event *k* at time *t* given *no other event k* or *k*^*′*^ has yet occurred. Furthermore, *h*_*k*_(*t*|*X*) can be modeled directly in a regression model with a proportional hazards specification, viz,

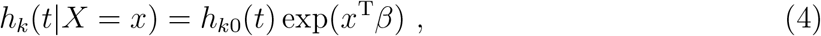

where the baseline sub-distribution *h*_*k*0_(*t*) is non-parametric. Fortunately, we do not need to be overly concerned with the interpretation of *β*. Rather, for given values of *x, β*, and *h*_*k*0_(*t*), we can compute—or estimate—the *h*_*k*_(*t*|*X* = *x*) and thereby obtain *F*_*k*_(*t*|*X* = *x*), and functions deriving therefrom including the complete sub-distribution CIF. Estimation of and inferences on *β* and *h*_*k*0_(*t*) are given by Fine (1999).

A final consideration relates to statistical inferences—hypothesis tests and confidence intervals. In the demonstration analysis in the next section, we did not obtain confidence intervals for estimated parameters; we were still adapting methods to the rapidly changing COVID-19 landscape. However, in ongoing work, for estimates such as the probability of ICU use, either globally or stratified by grouping variables such as age, we have typically employed the bootstrap procedure (Efron, 1994), sampling from the entire set of hospitalizations with replacement, and re-estimating the entire parameter set for each resample. We then obtain bootstrap standard errors and employ normal-theory-based tests and confidence intervals. Alternatively, for tests comparing groups with respect to sub-distribution hazard, we have employed likelihood ratio tests from the proportional hazards model (4), with sets of indicator variables representing the groups or strata.

## 3 Example: Hospital System Data in Austin, Texas

### 3.1 Study Population and Data

Data arise from one of three large hospital systems in the 5-county Austin-Round Rock Metropolitan Statistical Area (MSA, e.g., CDC, 2018), located in Central Texas and comprising Travis, Williamson, Bastrop, Caldwell, and Hays Counties. As the pandemic grew in March and April, 2020, we developed a system to extract data from the backend database of the electronic health record (EHR) system at use in the inpatient settings across the system, with the goal of capturing each COVID-19 hospitalization in the health care system. The data contain several key elements: patient id, which is valid across all hospitals in the system, date-time (to the nearest minute) of admission, occurrence and date-time of ICU admission, date-time of ICU discharge, date-times of initiation and termination of mechanical ventilation (Vent), date-time of hospital discharge, and whether the exiting event was a live discharge or in-hospital death. In addition to the above, the data include a vast array of demographic and clinical indicators available at admission (baseline).

This and subsequent studies using these data were approved by the University of Texas at Austin Human Subjects Institutional Review Board.

Regarding details of data acquisition, data were extracted from the EHR database using a custom SQL query on the hospital system (versus Dell Medical School) side, which pulls directly from an encounter table, where there is one row per encounter, and then joins to person level tables for person level demographics such as age, sex, race/ethnicity, etc. ICU data are pulled from a patient bed movement table and Vent data are pulled from an orders table. The query only pulls in the first and last instance of ICU and Vent admission/start and discharge/end times during an encounter, and left joins those to the encounter level data, so that any multiple ICU stays and/or uses of mechanical ventilation are considered as one long event. Comorbidity variables were pulled from a billing table in the EHR database. This means the comorbidity would only be present in the encounter if it was addressed and documented my the provider in such a way as picked up by billing coders.

Additional variable creation and tidying were done in a detailed R (R Core Team, 2019) script, sharable across our team across the hospital system and Dell Medical School. Inconsistencies in values for labs and vitals due to differences in provider documentation were resolved and variables with similar or redundant categories were condensed and simplified. Three important ones are mentioned here: First, only presenting labs/vitals are included. This was defined as the first measure with a date-time stamp of that lab/vital that occurred during the patient’s hospitalization. If that lab or vital is missing in the dataset, it was taken to mean the lab/vital was not measured during the hospitalization. Second, some facilities provide heart rate, whereas others document pulse. We combined these for definition of “presenting lab”, i.e., if a patient was missing heart rate, we then pulled in pulse; a similar approach was taken for temperature measured in F vs C. Finally, we collapse some discharge disposition categories such as “rehab” (for rehabilitation facility) and skilled nursing facility into as “Rehab/SNF”.

Data and analyses are presented here primarily to document and illustrate the competing risks methodology, and hence do not exploit most of these variables. In addition, key results used in the report by Yang et al. (2021) are included here. More in-depth, comprehensive, and up-to-date investigations are presented in companion reports, one of which (Rathouz, 2021) combines the data from the present hospital system with those from the other two major system in the Austin-Round Rock MSA. Other reports exploiting the detailed data collected here are forthcoming.

### 3.2 Data Organization for Analysis

Preliminary manipulation involved reconfiguring the data into a format of “one row per hospitalization”. We sought to identify transfers as pairs of hospitalizations that ended and started within 2 days of one another on the same patient at two different facilities. Aside from these combinations, a few patients did apparently have multiple hospitalizations. We treated these as independent observations.

Some minor adjustments were made to the data in the data cleaning phase. First, some individuals appeared to be admitted directly to the ICU; for those individuals, we dialed their hospital admission back by 30 minutes to be able to capture that event with standard survival analysis software tools, which require all events to have a positive time of occurrence. Similarly, in case of in-hospital death, we harmonized time of death and time of discharge by consolidating the two potential outcomes into a single “exit” time. For patients that expired, their exit time was set to their death time which is consistently recorded prior to their discharge time. For (very few) expired patients with missing death times, their exit was set to their discharge time. Remaining patients who were discharged normally had their exit time set to discharge time. Lastly, date-times for ICU and Vent that appear to occur following hospital discharge were also wound back to equal hospital discharge date-time to produce a logical ordering of hospitalization events for the patient from admission to discharge.

Data were then organized to have several key elements to conform to the statistical framework in Section 2.1. These include date-time of admission, an analysis indicator variable for whether ICU was employed, a date-time of ICU admission, date-time of ICU discharge, and a date-time of hospitalization discharge, along with an indicator as to whether that discharge was a live discharge or represented in-hospital death. Vent occurrence and date-times were handled similarly to ICU date-times. Here, the primary goal of the data organization was to configure the data as closely as possible to map onto event times described in Section 2.1, namely *t*_0_, *t*_ICU_, *t*_Vent_, *t*_Disch_, and *t*_Cens_, along with *D*_ICU_, *D*_Vent_, and *D*_Disch_, from which *D*_Disch|ICU_ can also be computed. Four primary hospitalization event endpoints were considered in the analyses presented here, which include both the estimated probability of each event occurring and features of the times to those events, *conditional* on such events occurring. These include:

- ICU use (*D*_ICU_ = 1) and time to ICU from admission, *t*_ICU_ − *t*_0_.
- discharge without (bypassing) ICU (*D*_ICU_ = 2) and time to discharge from admission given this event, *t*_Disch_ − *t*_0_.
- given ICU use, live discharge (*D*_Disch|ICU_ = 1) and, given such, time to discharge from ICU admission, *t*_Disch_ − *t*_ICU_.
- given ICU use, in-hospital mortality (*D*_Disch|ICU_ = 2) and, given such, time to death from ICU admission, *t*_Disch_ − *t*_ICU_.

Survival analyses were all performed in Stata (StataCorp, 2019).

### 3.3 Results

The data comprise hospitalizations on 766 girls/women and 872 boys/men (representing biological sex), with the following age distribution:

| 0–4 | 5–17 | 18–49 | 50–64 | 65+ | Total |
| --- | --- | --- | --- | --- | --- |
| 14 | 41 | 603 | 480 | 500 | 1638 |

Of the 1,638 admissions, 518 of those patients were also admitted to ICU, with 43 still in hospital but not in ICU at time of administrative censoring of data. Figure 1 presents cumulative admissions to the system for COVID-19 by date.

**Figure 1:**
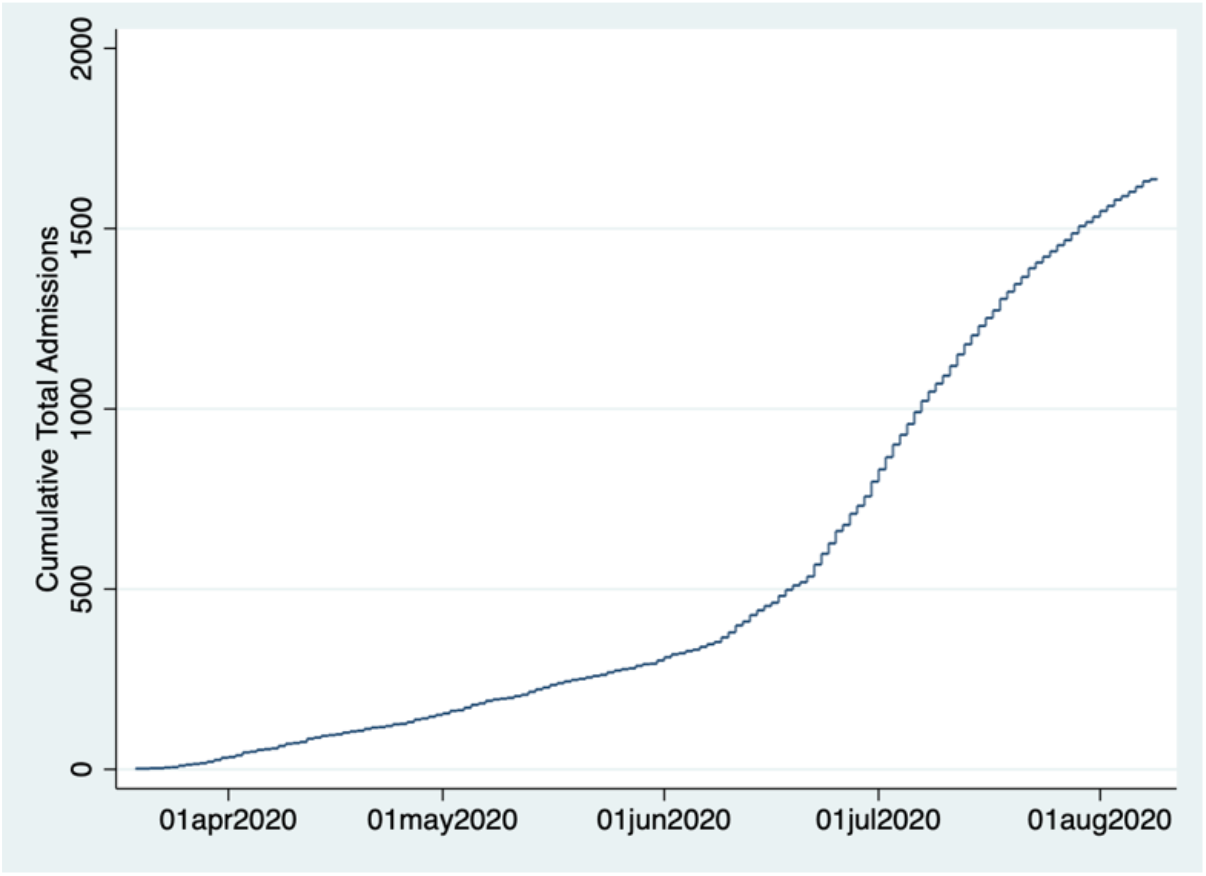
Cumulative system admissions by time. Cohort of COVID-19 hospitalized patients, single multi-hospital system, Austin, TX. Admits from 19 March through 08 August, 2020.

Tables 1 and 2, and Figure 2, present the probability of using (Table 2) or not using (Table 1) the ICU, along with features of the time to either ICU admission or hospital discharge *given* ICU use or not. As stated earlier, these parameters were chosen to support needed inputs to mathematical epidemic modeling of the local pandemic and pandemic response. Figure 2 presents the CIF’s for both entry into the ICU (red) and discharge from hospital bypassing ICU (blue). As can be seen, a very high proportion of ICU users are admitted to ICU essentially upon admission to hospital, but some are delayed or a day or two. Nevertheless, almost all users are admitted by about Day 5. Discharge without ICU use happens more gradually, with more than 90% of those being discharged, doing so by about Day 10. Note that the red and blue curves plateau at the estimated probability of either using ICU (0.32) or bypassing it (0.68).

**Table 1:**
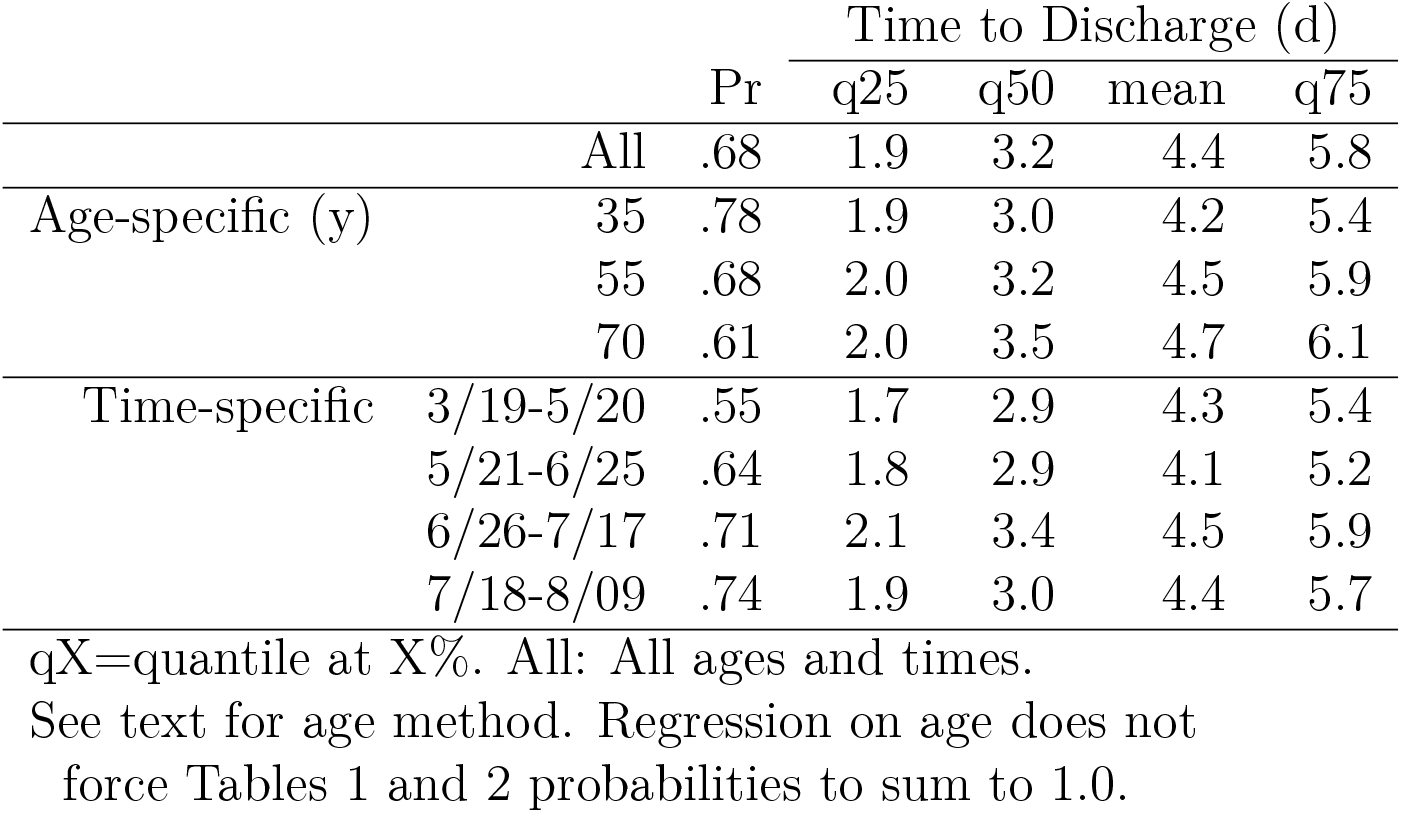
Probability of bypassing ICU and time to hospital discharge given no ICU. Cohort of COVID-19 hospitalized patients, single multi-hospital system, Austin, TX. Admits from 19 March through 08 August, 2020.

**Table 2:** Probability of ICU admission and time to ICU admission given ICU admission. Cohort of COVID-19 hospitalized patients, single multi-hospital system, Austin, TX. Admits from 19 March through 08 August, 2020.

|  |  | Pr | Time to ICU Admit (d) |  |  |  |
| --- | --- | --- | --- | --- | --- | --- |
|  |  |  | q25 | q50 | mean | q75 |
| All |  | .32 | .11 | .22 | 1.3 | 1.6 |
| Age-specific (y) | 35 | .26 | .11 | .23 | 1.3 | 1.7 |
|  | 55 | .32 | .11 | .22 | 1.3 | 1.6 |
|  | 70 | .36 | .10 | .21 | 1.2 | 1.6 |
| Time-specific | 3/19-5/20 | .45 | .13 | .19 | .76 | .76 |
|  | 5/21-6/25 | .35 | .12 | .23 | 1.0 | 1.1 |
|  | 6/26-7/17 | .28 | 0 | .22 | 1.5 | 2.0 |
|  | 7/18-8/09 | .24 | .07 | .24 | 1.8 | 2.3 |

p1=proportion in ICU by 1d.
qX=quantile at X%.
All: All ages and times.
See text for age and time methods.
Note: Additional parameters estimated for the entire cohort include the proportion being admitted to ICU within the first day: 0.68; and the time to admission given admission is >1d: 2.9d.

**Figure 2:**
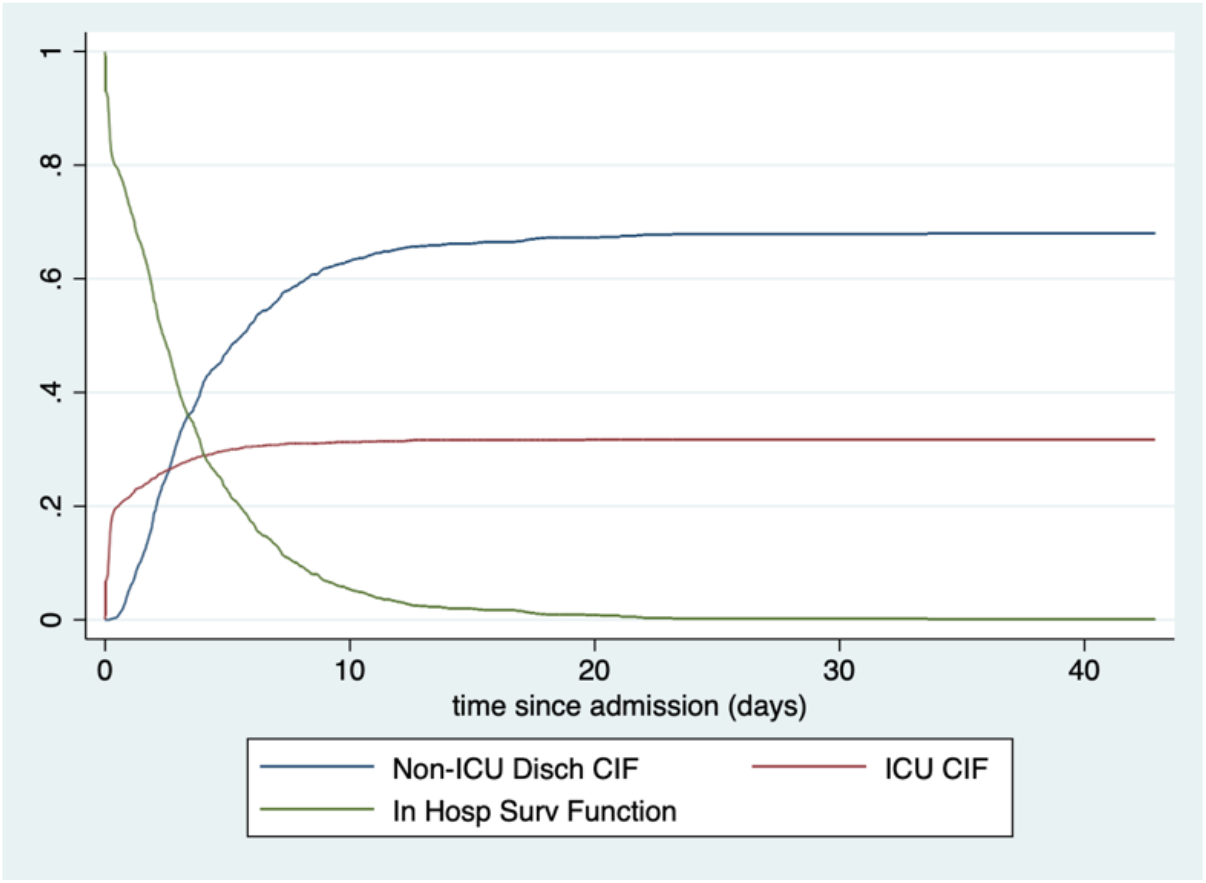
Estimated cumulative incidence functions for discharge without ICU use and for admission to ICU. Cohort of COVID-19 hospitalized patients, single multi-hospital system, Austin, TX. Admits from 19 March through 08 August, 2020.

We also present results by patient age and by calendar time-window in Tables 1 and 2. Without repeating the entire tables, main patterns are as follows: Probability of using ICU increases with age and decreases with calendar time. Time in hospital for non-ICU users increases with age.

Because focus here is on the methodological approach, we would like to point out how calendar time and patient age were handled differently. For calendar time, we stratified the data into 4 time periods based on admission date-time, and performed estimation on each period separately. For age, we fitted a Fine and Gray (1999) proportional hazards model for the sub-distribution hazard, and then extracted the fitted CIF at each of the three ages.

Tables 3 and 4 and Figure 3 present results only for those patients who are admitted to ICU. Times are time *since ICU* admission. From the red CIF in Figure 3, it can be seen that about 22% of COVID-19 patients who enter ICU die in the hospital, with a very high proportion of those dying by about 30 days. Probability of in-hospital mortality is strongly related to age, but time to death is very similar for all ages. For those who survive, length of stay is longer for older patients. Differential results by calendar time, given ICU admission, were not of interest for local mathematical epidemic models, so we did not generate results stratified by time.

**Table 3:** Probability of live hospital discharge and time from ICU admission to hospital discharge, given ICU admission ending in live discharge. Cohort of COVID-19 hospitalized patients, single multi-hospital system, Austin, TX. Admits from 19 March through 08 August, 2020.

|  |  | Time to Discharge (d) |  |  |  |  |
| --- | --- | --- | --- | --- | --- | --- |
|  |  | Pr | q25 | q50 | mean | q75 |
| All |  | .78 | 5.8 | 10.8 | 17 | 23 |
| Age-specific (y) | 35 | .91 | 5.0 | 9.1 | 14 | 17 |
|  | 55 | .80 | 6.0 | 11 | 17 | 24 |
|  | 70 | .70 | 6.7 | 12 | 19 | 28 |

qX=quantile at X%. All: All ages.
See text for age method. Regression on age does not force Tables 3 and 4 probabilities to sum to 1.0.

**Table 4:** Probability of in-hospital death and time from ICU admission to death, given ICU admission ending in in-hospital death. Cohort of COVID-19 hospitalized patients, single multi-hospital system, Austin, TX. Admits from 19 March through 08 August, 2020.

|  |  | Time to Death (d) |  |  |  |  |
| --- | --- | --- | --- | --- | --- | --- |
|  |  | Pr | q25 | q50 | mean | q75 |
| All |  | .22 | 4.6 | 10 | 12 | 19 |
| Age-specific (y) | 35 | .06 | 5.0 | 11 | 13 | 21 |
|  | 55 | .16 | 4.9 | 11 | 13 | 19 |
|  | 70 | .31 | 4.8 | 10 | 13 | 19 |

qX=quantile at X%. All: All ages.
See text for age method.

**Figure 3:**
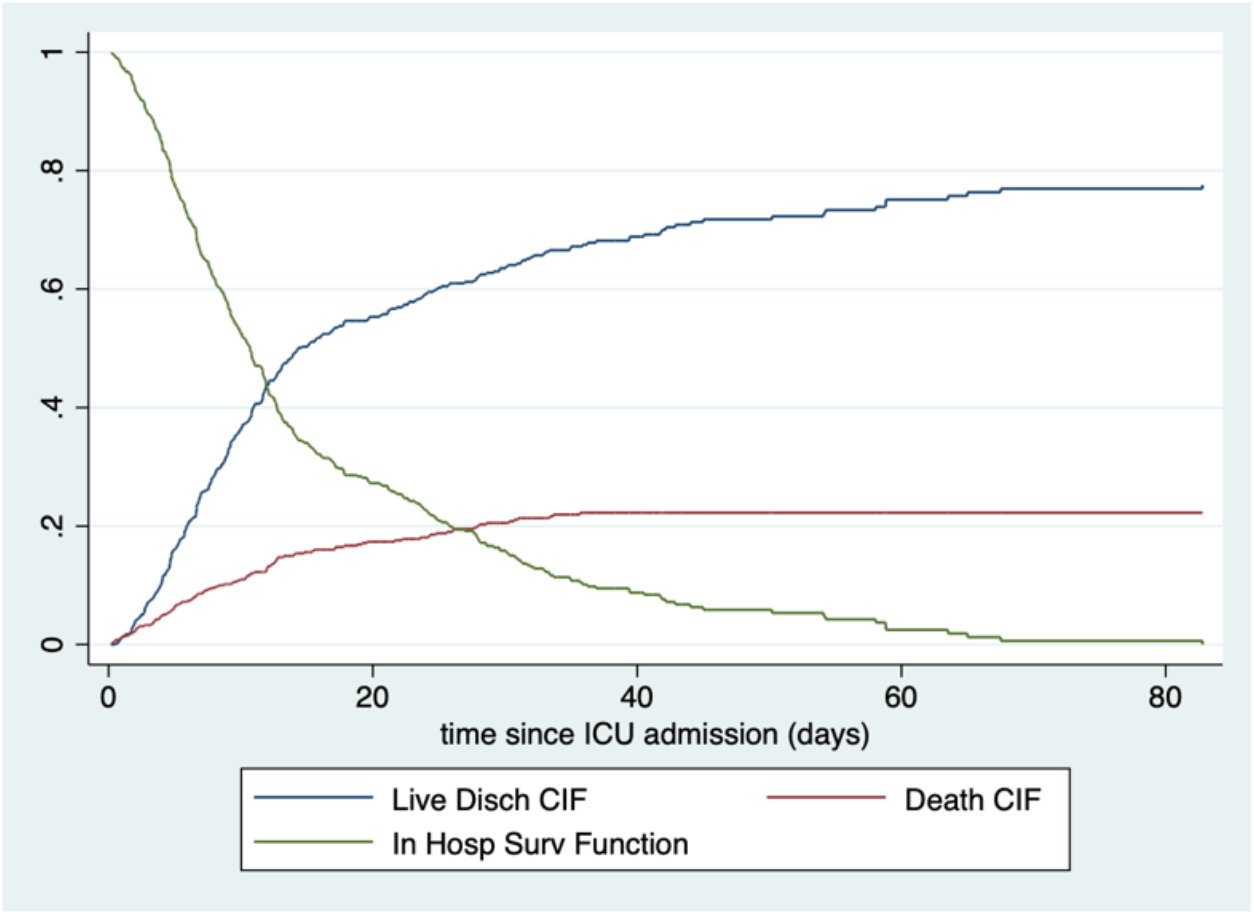
Among patients admitted to ICU, estimated cumulative incidence functions for time from ICU admisson to either live discharge or in hospital death. Cohort of COVID-19 hospitalized patients, single multi-hospital system, Austin, TX. Admits from 19 March through 08 August, 2020.

## 4 Discussion

Owing to the fact that separate models are fitted for each CIF, probabilities do not always sum to 1.0. We see this in the age-specific estimation in Tables 3 and 4. There is nothing forcing the two models for the age effects to be harmonized. This is an area for further investigation.

We used stratification to model the effect of calendar time, and regression to model that of age. The former is likely somewhat statistically inefficient, while the latter could be subject to bias if the effect of age is not linear on the log-hazard scale and/or the reference hazard function does not hold across the entire age range. A compromise approach would be to model age in the regression part of the model more flexibly, possibly using indicator variables for different ranges, while still maintaining the proportional hazards assumption (i.e., still assuming that that all the age strata share a common reference hazard function).

A related issue is the subtle interpretation of the proportional sub-distribution hazards model (4). We believe we have now located a more interpretable model for the effect of predictors and covariates based on the logistic transformation (Eriksson et al., 2015), which yields parameters on the traditional log-odds scale. This will be exploited and illustrated in future reports.

## Data Availability

These data are not available for distribution.

